# Viral Mutations as a Possible Mechanism of Hidden Immunization and Containment of a Pandemia

**DOI:** 10.1101/2020.04.09.20059782

**Authors:** Juri Dimaschko, Victor Podolsky

## Abstract

An impact of viral mutations on the extent of an epidemic is examined. A mechanism of immunization of the population via spread of weakly mutated strain as a natural factor terminating the epidemic is indicated. An epidemic model which details this mechanism is proposed.

## I. INTRODUCTION AND MOTIVATION

The COVID-19 virus has spread across the world, becoming a pandemic. It brings with it a great deal of social, economic, and political damage.

In this rapidly developing crisis, it is necessary to reliably assess the current state of the epidemic as well as to predict its near- and medium-term development. A scientifically sound assessment is vital for deciding on how to allocate the significant yet limited medical resources available to combat the epidemic. And it is crucially important for determination of optimal quarantine measures, such that they would effectively reduce the severity of the epidemic but would not inflict near-irreparable damage to the economy.

In this paper, we consider the role of the inherent mutability of the virus. We recognize it as the principal factor which critically suppresses the growth of novel viral epidemics. The mechanism of this suppression has a well known evolutionary nature. In plain words, once viruses get under attack of the immune systems of the infected humans, they mutate to survive in the hostile environment. Given the diminutive scale of the virus reproduction time, it is fair to assume that viruses may mutate quite fast on the human timescale. And by evolutionary principle, a less pathogenic virus strain has better survival chances. When a statistically significant number of people get infected, there should be a large number of them with strong immunity that facilitates less pathogenic virus mutations. As these people transmit their weaken strain to others, we observe some natural cross-immunisation.

We develop a new mathematical model which takes viral mutability into account, and compares our results against other models and current factual data available for COVID-19 pandemic. Our results allow us to evaluate our ability to suppress the spread of the epidemic, and to debate on measures which we should take in order to optimally facilitate its containment and termination. We also provide a medium term forecast of the epidemic development in those countries where COVID-2019 has yet to unravel in full force.

The paper is structured as follows.

In the first part we review the commonly accepted SIR epidemiological model [1]. We argue that the SIR-based estimates of the peak values of the infected population and epidemic duration can hardly be supported by COVID-19 data for a number of countries. We attribute this fact to the SIR model overlooking the impact of the virus mutability on the epidemic, whereas the fact of rapid mutations is now reliably established [2].

We next extend the SIR model to incorporate a virus mutation factor. Our model, which we call SIMR, shows that rapid virus mutations that accompany avalanche epidemic phase can drastically scale down the epidemic and reduce the height of its peak by a factor of ten-to-hundred as compared to the SIR results. In the second part of this paper, we use the SIMR model to analyze the now practically terminated COVID-19 outbreak in Wuhan of China [3]. This allows us to evaluate the main internal parameter of the model - the probability of mutation of the transmitted virus. Next, we shall use this approach to analyze and assess the current situation in other countries where the epidemic still goes.

In the third part, we analyze the current state of the epidemic in a number of countries and megacities, as well as make medium-term forecasts of the development of the situation. The analysis is based on the same SIMR model with the same basic parameter.

In the conclusion, we qualitatively evaluate the role of the SIMR model parameters in limiting the scale of the epidemic as well as discuss the role of the age structure of a country population which is not explicitly reflected in the SIMR model in the suggested form.

## II. TWO MODELS OF THE EPIDEMIC DEVELOPMENT

### A. Basic SIR model that does not take into account mutations of the virus

It is the main epidemic development model that exists today to describe the course of an epidemic. It is based on a three-stage scheme, where the entire population is divided into three parts: *S* is susceptible (uninfected), *I* is infected (infected, sick), *R* is recovered.

It is assumed that the survivors acquired perfect immunity at the cost of the disease and no longer get sick. Taking into account that *S*(*t*), *I*(*t*) and *R*(*t*) are the fractions of these groups throughout the entire population, the model builds the evolution of the epidemic through a system of three differential equations:

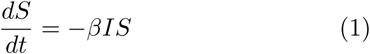

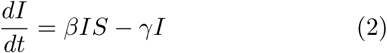

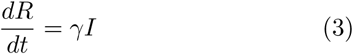

The model parameter *β* characterizes the rate of transmission of the virus, and *γ* the rate of recovery and thereby the acquisition of immunity. The SIR model keeps the sum *S*(*t*) + *I*(*t*) + *R*(*t*) = 1 constant, as it should be.

Point *S* = 1, *I* = *R* = 0 corresponds to a state with no infected. Small deviations from it develop in time according to the law *∝* exp [(*β – γ*) *t*]. The emergence of the epidemic and its further development is controlled in this model by the only dimensionless parameter *r* = *β/γ*. At a high propagation speed of the virus, when *r* is more than one, the number of sick people starts to grow and the epidemic begins.

Later on, the number of sick people increases until a significant majority of the population passes through the stage of the disease and thereby acquires immunity. At the beginning of the epidemic, the vast majority of people *S*(0) = 1 – ≪ *I*_0_ are healthy *I*(0) = *I*_0_ 1 is the initial fraction of carriers of the virus, *R*(0) = 0 - nobody have been sick and gained immunity so far). During the epidemic, a significant fraction of people 0 *< I* (*t*) *<* 1are sick. At the end of the epidemic, there are no more sick people, *I*(*t*) = 0. Some people *R*(*t*) as a result of the epidemic went through disease and gain immunity, another part *S*(*t*) escaped the disease. The typical course of the epidemic in the SIR model is as follows (Fig.1): The maximum fraction of the sick people during the epidemic is given by the relation [1]

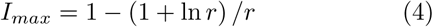

**FIG. 1.**
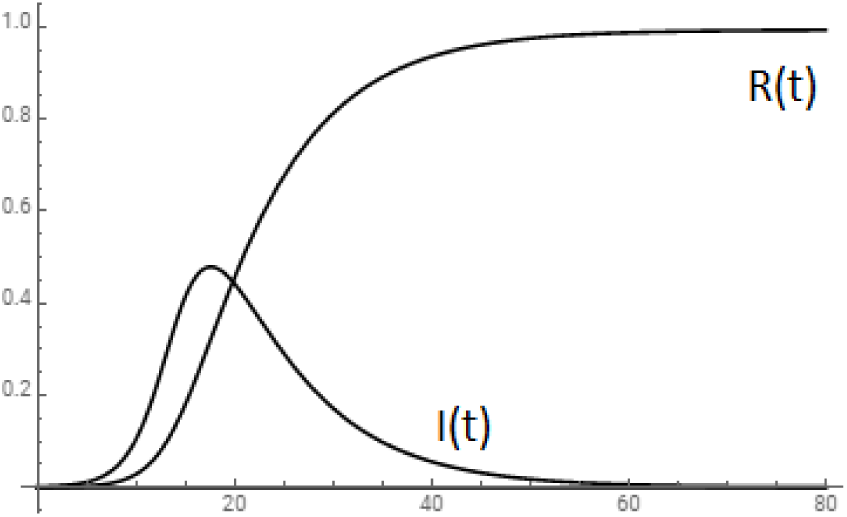
SIR model epidemic development (*β*=0.6, *γ*=0.12).

It follows from this that the fraction of sick people at the peak of the epidemic can be very high. So, at *r* = 5 it is *I*_*max*_ = 48% as shows the I(t) plot in Fig.1. Experience shows that the fraction of sick people never reaches this level. This fact alone indicates the incompleteness of the SIR model and the existence of other mechanisms for ending the epidemic, not related to the acquisition of immunity by people through the stage of the disease.

### B. New SIMR model taking into account virus mutations

To understand what exactly can be this way, you need to consider that the virus itself, like the human body, changes during the course of the epidemic. And if a person gains immunity in the course of a disease, then the initial strain of the virus, due to the sharply increased replication rate in the body affected by the disease, rapidly mutates.

Mutations of the virus occur randomly, but it is precisely those strains that lead not to acute, but to a subnormal, latent course of influenza that are transmitted most successfully. It is the carriers of such a strain that transmit the largest number of viruses, and therefore it is precisely such a strain that spreads most rapidly and most successfully. Thereby there is a kind of mutual complementarity:

1. **an aggressive strain** quickly and efficiently propagates inside the body affected by the disease, but it has a limited duration of action, during which it either is eliminated by the immune system or leads to the death of the body. Further, he is less likely to spread further due to the rapid immobilization of the patient;
2. **a milder strain** does not lead to acute illness and multiplies inside the human body to a much lesser extent, but it has the ability to remain in the body for a long time, because it is built into the subnormal state of the body. Further, he is more likely to transmit from person to person due to the lack of external manifestations of the disease.

For these reasons, the softer strain spreads faster and thereby exhausts the full area of the virus, gradually depriving the original aggressive strain of the possibility of further spread. In general, this should lead to a change in the composition of the strain circulating during the epidemic in the direction of its mitigation.

Further, although the mutated virus does not lead to an acute course of the disease, the very scale of its reproduction in the human body is limited by the activation of the natural immune mechanism. Due to the genetic proximity of the original and mutated viruses, the effect of the latter acts as a natural vaccine, leading to hidden immunization of the body.

Both of these factors the softening of the initial strain and the hidden immunization act in the direction of ending the epidemic.

Virus mutations can be included in the base SIR model, giving the original virus ability to mutate during transmission.

For this, along with the previous three categories (susceptible - infected - recovered), the fourth is introduced - those who received the mutated virus. The fraction of such people who got a mutated virus and were protected by it from the disease is denoted by *M* (*t*). In the future, they do not get sick and live the same way as they were recovered persons from the set *R*(*t*).

Let the probability of mutation in a single act of virus transmission be *m <* 1. Thus, the fraction of sick people I(t) will be replenished at a rate of *β* (1 – *m*) *IS*. The fraction of people infected with a mutated virus *M* (*t*) will be replenished at a rate of *βmIS* from sick people and at a rate of *βMS* from those who already got mutant virus. Then equations of the modified model, similar to the base one, take the form

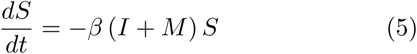

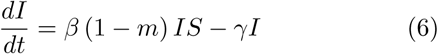

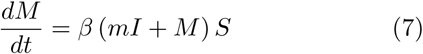

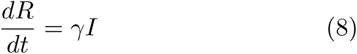

Later on, we will call it the SIMR model. At *m* = 0, it leads to the absence of mutations, as a result to *M* (*t*) = 0, and just coincides with the basic SIR model.

In the essence, the SIMR model is based on the model assumption that there are only two strains of the virus - the original (aggressive) and mutated (milder) ones. Now it is important to understand the effect of the mutation factor *m* on the epidemic in this model.

Like the SIR model, the SIMR model keeps the sum *S*(*t*) + *I*(*t*) + *M* (*t*) + *R*(*t*) = 1 constant. Point *S* = 1, *I* = *M* = *R* = 0 corresponds to the state with no infected people. Small deviations from it develop in time according to the law *∝* exp [(1 – *m*) (*β – γ*) *t*]. The emergence of the epidemic and its further development is controlled in the SIMR model by the only dimensionless parameter *r* = (1 – *m*) *β/γ*. At a sufficiently high rate of spread of the virus *β*, when *r >* 1, the fraction of sick people begins to grow and an epidemic occurs.

A typical course of the development of the epidemic when mutations are taken into account is presented in Fig.2.

A characteristic feature of the mutation model is a significantly smaller fraction of sick people. This is due to the fact that the vast majority of people got a one and thereby undergo the hidden immunization.

On the graph, this is manifested in the fact that during the course of the epidemic, the fraction of *M* (*t*) who got a new, mutated virus significantly exceeds the fraction of *R*(*t*) who got the original strain and thereby got sick.

We emphasize that from the point of view of epidemiology, these two groups are equivalent - both of them are no longer sick and gained immunity. Although the price they paid for it is different. The first group (M) suffered the disease subnormally and barely noticed anything. The second group (R) suffered the disease in severe form with all the attendant risks.

Since a person got a mutated virus does exhibit any pronounced disease and practically does not appear externally, it is natural to interpret the value of *M* (*t*) as a percentage of **hidden immunization**.

**The key effect manifested in the SIMR model of the epidemic is the avalanche-like spread of the mutated strain, which is significantly faster than the epidemic itself. As can be seen from Fig.2, at the peak of the epidemic, the percentage of the hidden immunization** *M* (*t*) **caused by this strain already exceeds 70%, and soon afterwards tends to 100%. In the framework of the SIMR model, this is precisely the terminating factor of the epidemic**.

Thus, the SIMR model describes a natural mechanism for limiting the scale of the epidemic, which is based on a mutation of the original virus, its gradual displacement from the circulating strain, and hidden immunization of the population with the mutated strain.

To assess the level of the hidden immunization, it is convenient to consider the limiting case of the absence of the usual immunity, *γ* = 0. In this case, during the course of the epidemic, all susceptible persons sooner or later get either the original strain - *I*(*t*) - or the softened mutated one - *M* (*t*). In this limit, the SIMR model has an exact solution (see Appendix A), allowing to find the limiting value of the fraction of infected *I*(*t*) by the end of the epidemic, i.e. as *t* → ∞:

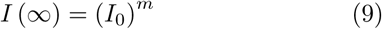

For example, with an initial infection rate of one in a million, *I*_0_ = 10^*-*6^, and *m* = 0.2, the total infection rate of the original strain is *I* (∞) = 1*/*16. This is part of those who have been immunized through the disease. The rest will receive a mutated virus, *M* (∞) = 1 − *I* (∞) = 15*/*16, or 93.8%. This is the fraction of immunity resulting from the epidemic through hidden immunization. At *m* = 0.4, the total fraction of the hidden immunization increases to 255/256, i.e. up to 99.6%.

It is clear that Eq.(9) gives the upper limit of the maximum value of *I*(*t*) because the recovery term in Eq.(2) is negative.

These examples show a strong influence of the probability of mutation m on the fraction of those who had the initial strain during the epidemic and for any nonzero *γ* by the end of the epidemic there are no sick people, *I* (∞) = 0.

The used approximation *γ* = 0 is good to describe the rise of epidemic, since in this stage spread of the virus dominates. In this approximation the SIMR model has exact solution **for any dependence** *β*(*t*). This is important since such dependence as well as the initial spread rates *β*(0) are unknown. They are affected by traditional modus vivendi of people in different countries and by different quarantine measures, and it is very hardly to evaluate both factors in a reasonable way.

Fortunatelly it turns out to be possible to exclude the dependence *β*(*t*) from the result and to connect directly two main values, *M* and *I*, in a simple relation (see Appendix A):

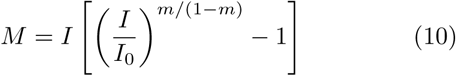

**Thereby one can trace the invisible level of the hidden immunization** *M* **as the fraction of sick people** *I* **is known at any time**. Further we find the mutation factor *m* from the data of Wuhani epidemic and then will be in a position to evaluate the current level of the hidden immunization in different countries, where the epidemic still is rising. Eq.(10) gives the lower limit of the current value of *M*. This can be seen from the end limit *t* → ∞ when *I* = 0 and *M >* 0.

In the point of maximum the right side of Eq.(6) is zero, that immediately gives *S* = 1*/r*. As *M ≤* 1 – *S*, this means

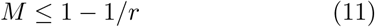

Thereby in rise of the epidemic we have the lower limit (10), and in the maximum both lower and upper limits (10,11).

## III. SIMR SIMULATION OF THE WUHANI EPIDEMIC (CHINA)

The SIMR model has two main parameters.

The first of these is the rate *β* of spread of the virus. This parameter, being large enough, triggers the epidemic mechanism. For this, similarly to the basic SIR model, the condition must now be satisfied for a slightly different dimensionless parameter *r* = (1 – *m*) *β/γ*: *r >* 1. Here 1*/γ* is the characteristic time course of the disease. The virus spread rate *β* also determines the duration of the epidemic. In a wide range of other parameters, the time for the epidemic to reach a maximum is approximately 10 cycles, the duration of each of which is the time of a single act of virus transmission, 1*/β*.

The second parameter of the SIMR model is the probability of mutation m during virus transmission. The main property of this parameter is its determining effect on the maximum level of the epidemic, i.e. on the number of cases at its peak. This number rapidly decreases with increasing m according to the law (1 – *m*)^10^, which corresponds to a decrease in the fraction of the initial strain with factor(1 – *m*) for each cycle of virus transmission and its replacement with a softened, mutated virus.

The parameters *β* and *m* should be gained from combining statistics with the prediction of the SIMR model. In the case of interest to us, the most complete data are available on the epidemic of the COVID-19 virus in Wuhan (China), where it originated and is already close to completion. We will take as a basis the evolution of the current number of sick people for the entire period, and overlay the theoretical graph of the SIMR model on these data:

The code for solving the equations of the SIMR model, giving the theoretical The coincidence shown is achieved with a virus transmission time of 1*/β* = 2.2 days and a typical disease progression time of 1*/γ* = 14 days.

The probability of the viral mutation during its transmission based on these data is *m* = 0.39. Later on, we consider this value a constant, which is an internal property of the virus.

The dimensionless epidemic factor from here is *r* = (1 – *m*) *β/γ* = 3.8. Thus, the epidemic condition *r >* 1 is fulfilled, as it should be.

The irreparable discrepancy between the theoretical curve and statistics in the vicinity of the maximum, between the 15th and 30th days of the epidemic, allows for a fairly simple explanation: it was during this period that the Chinese authorities took unprecedented quarantine measures at the center of the epidemic, which of course reduced the virus transmission rate *β* by several times. In the SIMR model with a constant parameter *β*, this is not explicitly taken into account, but the qualitative effect of these measures on the dependence *I*(*t*) is obvious. From a comparison of the two lines on the graph, the effect of restrictive measures is visible. They lowered the maximum number of cases by about 25,000, which, with a 6 percent mortality rate, is about 1,500 saved lives.

Further, we can compare the maximum percentage of cases in Wuhan (with a population of 12 million, this is about 0.5%) with the prediction of a basic SIR model that ignores virus mutations. According to the above formula (4), for *r* = *β/γ* = 6.3, this value should be 55%, which is completely unrealistic.

It is also extremely important that the SIMR model allows us to track the growth of the percentage of hidden immunization M(t), which is not yet available to direct measurements. According to the SIMR model, at the peak of the Wuhan epidemic (on its 25th day) it was already 80%, on the 40th day 99.98%, and now, on the 80th day, i.e. on 11/02/2020, is almost 100%. This is what allows us to consider the Wuhan epidemic to be over.

## IV. SIMR-MODELING AND FORECAST OF THE COVID-19 EPIDEMIC IN SEVERAL COUNTRIES

Next, we use the SIMR model to analyze the current state of the epidemic in some specific cases. We note right away that the model proceeds from constant rather than local parameters, and therefore can pretend to describe only the average distribution of the epidemic in each country, developing only in time. At the same time, it is clear that in megacities and conglomerates it should develop much faster. This is exactly what is observed during the development of the pandemic. In order to smooth out this discrepancy between the model and the real situation, we will choose in each country the reference region of compact residence, which accounts for the majority of infections. For example, in Italy such a region is Lombardy with a population of 10 million people (of the total population of Italy 60 million). The following is a summary table of such reference areas / megacities for various countries to which the SIMR model will be applied (see Tab.1).

**TABLE I.**
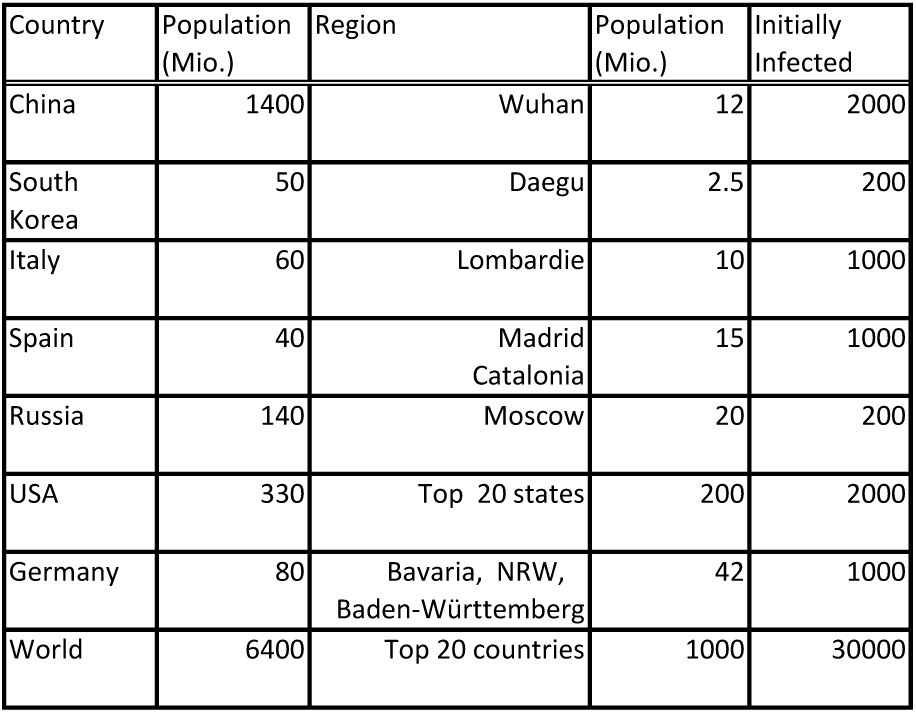
Reference regions

The initial number of infected in each case is selected from the principle of correspondence between the model and statistics. It has a logarithmically weak influence on the result, however, it is given here in order to enable complete verification of the model calculations. Further, it is understood that the rate *β* of the virus spread during an epidemic can and should be reduced by restrictive measures. For this reason, the growth in the number of infected people is slowing. Thus, a SIMR model with a constant value of the parameter *β* can only show a pessimistic assessment of the course of the epidemic, corresponding to the absence of dependence *β*(*t*). The initial value of the parameter *β* will be taken from the initial exponential portion of the observed dependence *I*(*t*) in the first 10-15 days of the epidemic. To increase the reliability of this assessment, we use the data starting from the moment when the number of sick people exceeds 1000.

We assume that the probability of a virus mutation is its internal property, does not depend on factors of external influence, and therefore can be directly taken from the Wuhan data in all cases: m=0.39. The time of exit from the disease is on average constant, determined by the average level of immunity and is 12-14 days. This corresponds to the rate value is about *γ*=0.07 and will be a bit varied for different countries for best fitting. The exception when this parameter is noticeably smaller is Italy, which have a large percentage of the elderly population with weakened immunity due to age. Thus, the expression for the increment of the dependence *I*(*t*), having the form of *α* = (1 – *m*) *β – γ*, allows to find the initial spread rate of the virus *β* from the established values of *α,γ*, and *m*.

### A. Italy

Here the epidemic develops according to the most dramatic scenario. The number of cases has already reached 80 thousand, and due to the large number of elderly among the cases, the mortality rate is noticeably higher than in Wuhan. The dynamics of the number of cases in Italy is shown in Fig. 4.

**FIG. 2.**
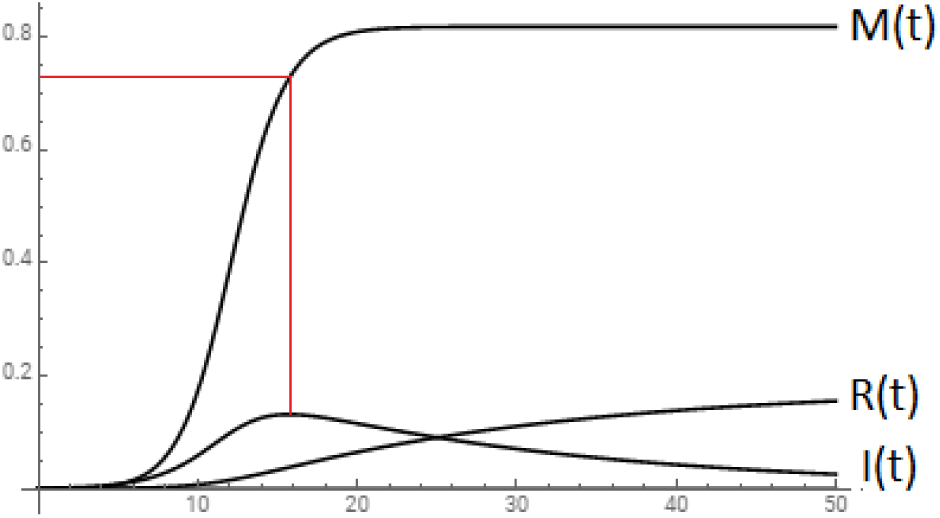
SIMR model epidemic development (*m*=0.2, *β*=0.6, *γ*=0.05).

**FIG. 3.**
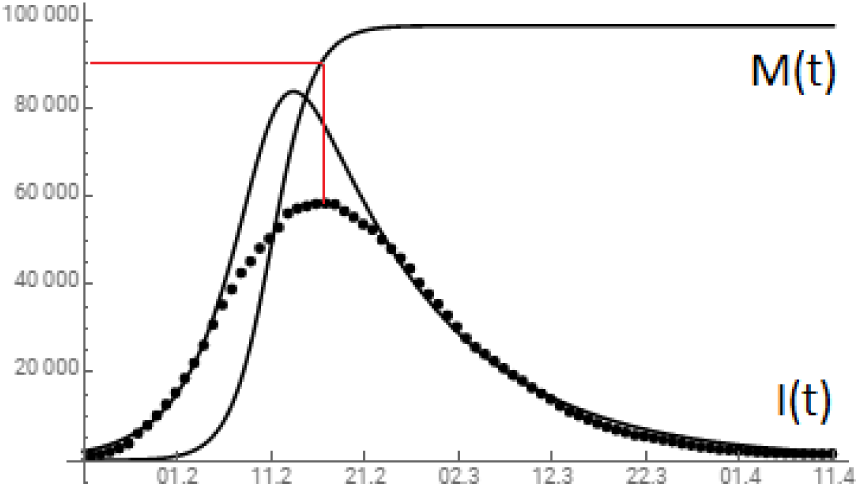
Comparison of the dynamics of the number of cases during the COVID-19 epidemic in Wuhan (China) according to the SIMR model (solid line) with statistical data [3] (dots). The SIMR parameters are taken as *m*=0.39, *β*=0.45, *γ*=0.071. Here and in all further figures, the line *I*(*t*) shows the number of cases, and the line *M* (*t*) is normalized to saturation with the value of *M* = 1 (100% hidden immunization). The red line indicates the hidden immunization *M* = 0.9 at the peak of *I*(*t*).

**FIG. 4.**
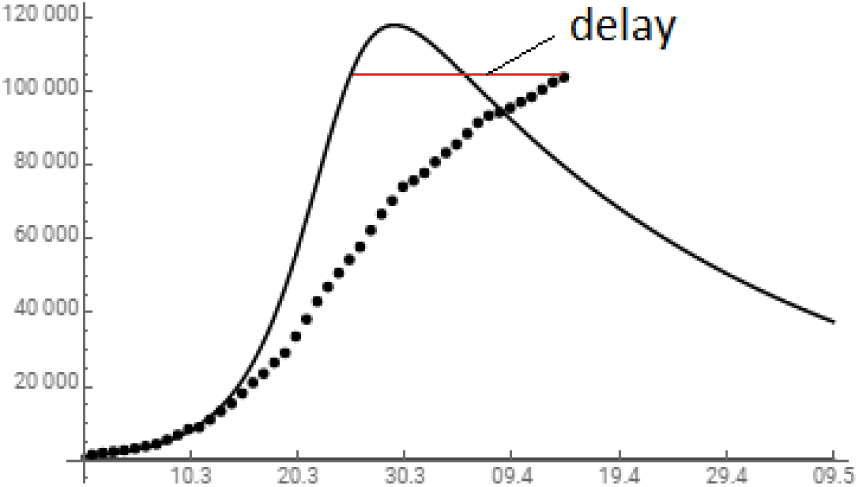
Comparison of the dynamics of the number of cases during the COVID-19 epidemic in Italy according to the SIMR model (solid line) with statistical data [3] (dots). The SIMR parameters are taken as *m*=0.39, *β*=0.39, *γ*=0.03.

The initial value of the increment is obtained from the number of sick people on 02.29.2020, *I*(0) = 1049 and after 10 days - on 10.03.2020, *I*(10) = 8514 [3]. Hence, the initial increment is 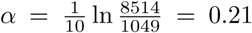. Given the known probability of mutation m=0.39 and a very small value of the parameter for the rate of exit from the disease *γ* = 0.03, this, in accordance with the relation *α* = (1 *-m*) *β - γ*, gives an estimate of the initial virus spread rate at the start of the epidemic: 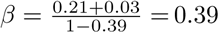. It is this value of *β* that leads to the model graph in the Fig.4.

The dimensionless epidemic factor from here is *r* = (1 *-m*) *β/γ* = 7.9. Thus, the epidemic condition *r >* 1 is fulfilled, as it should be.

It is clearly seen that after the first 2 weeks of the epidemic, the growth in the number of sick people slowed noticeably compared to the SIMR model with a constant value of *β*. It is natural to consider this as a consequence of the restrictive measures taken, leading to the decrease of *β*.

A comparison of the theoretical and observed dependences *I*(*t*) shows a noticeable lag of the latter by about 14 days. This indicates a later onset of the peak compared with the case of constant *β*, and the delay scale has already been determined, although it may slightly increase. Further, it is natural to assume that restrictive measures can only reduce the height of the maximum *I*(*t*), which for the theoretical curve is at the level of 120 thousand.

The lower limit of hidden immunization in Italy in accordance with Eq.(10) is about 20%.

Thus, in this example, we see that the SIMR model is not able to reproduce the exact course of the epidemic dependence *I*(*t*), but it gives an obvious upper limit on the height of the maximum and a reasonable estimate of the peak time. In this case, we are talking about another two weeks.

### B. Spain

Here the situation is similar to Italy. The number of cases exceeded 80 thousand and continues to grow. The dynamics of the number of sick people in Spain is presented in Fig. 5.

**FIG. 5.**
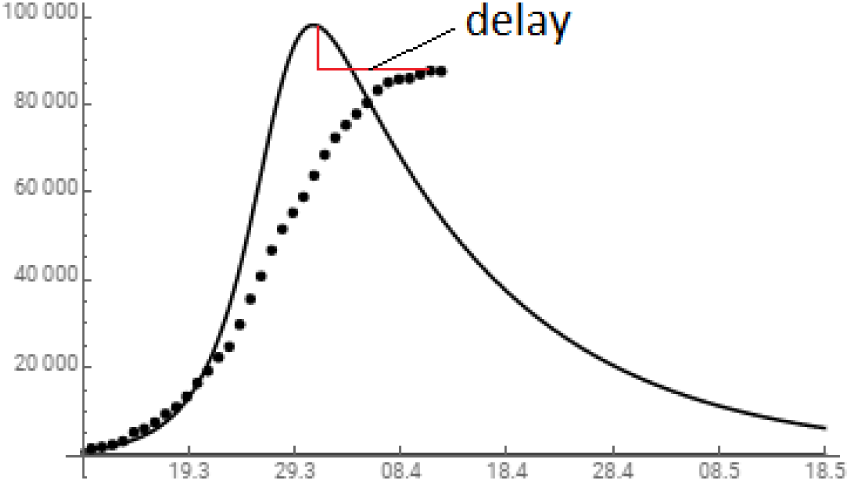
Comparison of the dynamics of the number of cases during the COVID-19 epidemic in Spain according to the SIMR model (solid line) with statistical data [3] (dots). The SIMR parameters are taken as *m*=0.39, *β*=0.52, *γ*=0.06.

The initial value of the increment is obtained from the number of sick people on 09.03.2020,*I*(0) = 1169 and after 10 days - on 19.03.2020, *I*(10) = 16139 [3]. Hence, the initial increment is 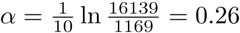. Given the known probability of mutation *m*=0.39 and a value of the parameter for the rate of exit from the disease *γ* = 0.06, this, in accordance with the relation *α* = (1 – *m*) *β – γ*, gives an estimate of the initial virus spread rate at the start of the epidemic: 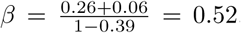. It is this value of that leads to the model graph in the Fig.5.

The dimensionless epidemic factor from here is *r* = (1 – *m*) *β/γ* = 5.3. Thus, the epidemic condition *r >* 1 is fulfilled, as it should be.

It is clearly seen that after the first week of the epidemic, the growth in the number of sick people slowed noticeably compared to the SIMR model with a constant value of *β*. It is natural to consider this as a consequence of the restrictive measures taken, leading to the decrease of *β*.

The lower limit of hidden immunization in Spain in accordance with Eq.(10) is about 9%. On the other hand, closeness to the maximum implies by Eq.(11) the upper limit of the hidden immunization as already about 80%.

A comparison of the theoretical and observed dependences *I*(*t*) shows a noticeable lag of the latter by about 10 days. This indicates a later onset of the peak compared with the case of constant *β*, and the delay scale has already been determined, although it may slightly increase. Further, it is natural to assume that restrictive measures can only reduce the height of the maximum *I*(*t*), which for the theoretical curve is at the level of 100 thousand.

### C. Germany

In Germany the epidemic is developing much slower than in other European countries and in the USA. The number of sick people here has already reached a maximum of 70 thousand and has stopped growing. The dynamics of changes in the number of cases in Germany is presented in Fig. 6. The initial value of the increment is obtained from the number of sick people on 08.03.2020, *I*(0) = 1022 and after 10 days – on 18.03.2020, *I*(10) = 12194 [3]. Hence, the initial increment is 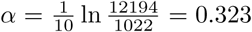. Given the known probability of mutation *m*=0.39 and a value of the parameter for the rate of exit from the disease *γ* = 0.075, this, in accordance with the relation *α* = (1 – *m*) *β – γ*, gives an estimate of the initial virus spread rate at the start of the epidemic: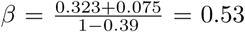. It is this value of that leads to the model graph in the Fig.6.

**FIG. 6.**
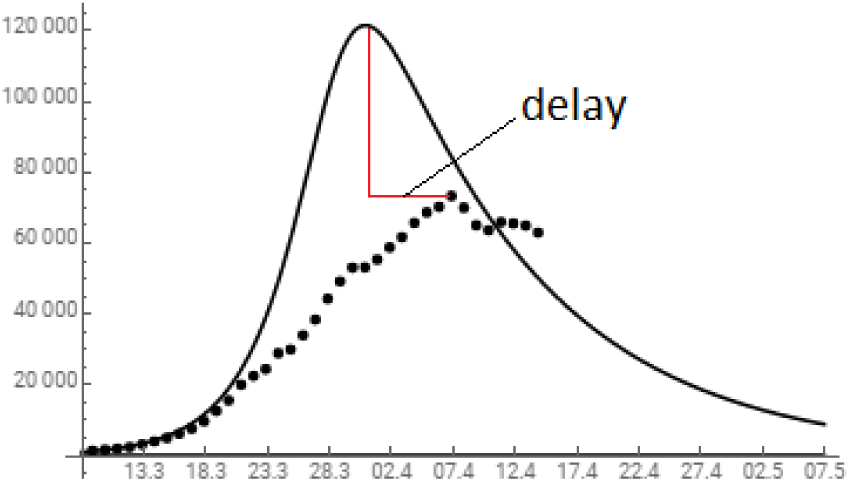
Comparison of the dynamics of the number of cases during the COVID-19 epidemic in Germany according to the SIMR model (solid line) with statistical data [3] (dots). The SIMR parameters are taken as *m*=0.39, *β*=0.53, *γ*=0.075.

The dimensionless epidemic factor from here is *r* = (1 *-m*) *β/γ* = 4.3. Thus, the epidemic condition *r >* 1 is fulfilled, as it should be.

It is clearly seen that after the first week of the epidemic, the growth in the number of sick people slowed noticeably compared to the SIMR model with a constant value of *β*. It is natural to consider this as a consequence of the restrictive measures taken, leading to the decrease of *β*.

As the epidemic in Germany passed maximum, by Eq.(11) the upper limit of the hidden immunization is currently about 75%.

A comparison of the theoretical and observed dependences *I*(*t*) shows a decrease of the maximum by about 50 thousand and it delay about 6 days. Both effects are an obvious consequence of the severe restrictive measures taken at the beginning of the epidemic. The flip side of such an effective restrictive approach is, from the point of view of the SIMR model, a certain slowdown in the growth of the hidden immunization and thereby delaying the maximum point of the epidemic - and hence its cessation.

However, as the epidemic maximum is already reached, the hidden immunization at this point is about 75%. This indicates the end of the epidemic within the next 10 days.

### D. Russia

In Russia the epidemic started much later as compared with other European countries and in the USA. From this reason the number of sick people is not high (about 15000) but grows rapidly. The dynamics of changes in the number of cases in Russia is presented in Fig. 7 The initial value of the increment is obtained from the number of sick people on 29.03.2020, *I*(0) = 1462 and after 15 days - on 13.04.2020, *I*(10) = 16710 [3]. Hence, the initial increment is 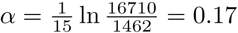. Given the known probability of mutation *m*=0.39 and a value of the parameter for the rate of exit from the disease *γ* = 0.05, this, in accordance with the relation *α* = (1 – *m*) *β – γ*, gives an estimate of the initial virus spread rate at the start of the epidemic: 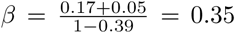. It is this value of *β* that leads to the model graph in the Fig.7.

**FIG. 7.**
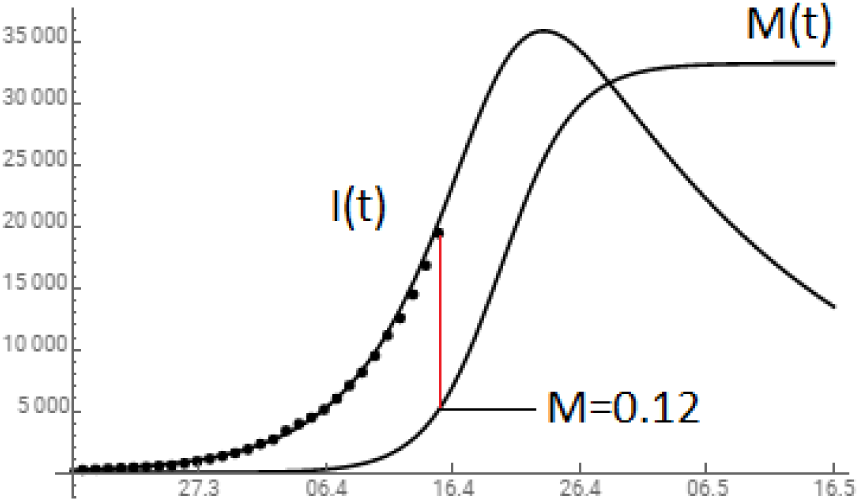
Comparison of the dynamics of the number of cases during the COVID-19 epidemic in Russia according to the SIMR model (solid line) with statistical data [3] (dots). The SIMR parameters are taken as *m*=0.39, *β*=0.35, *γ*=0.05.

The dimensionless epidemic factor from here is *r* = (1 – *m*) *β/γ* = 4.3. Thus, the epidemic condition *r >* 1 is fulfilled, as it should be.

Coincidence of the SIMR graph with statistical data enables to use the solution for M(t) as the value of the current hidden immunization in Russia. It is about 12%.

The epidemic in Russia in still in development and exhibits very good coincidence with forecast of the SIMR model, with no delay. The pick is expected in 10 days on the level of 35,000 sick people

### E. USA

In USA the epidemic is developing faster than in China and Europa, the number of cases has already exceeded 300 thousand and continues to grow. The dynamics of changes in the number of sick people in the USA is presented in Fig. 8. The initial value of the increment is obtained from the number of sick people on 13.03.2020, *I*(0) = 2126 and after 14 days - on 27.03.2020, *I*(14) = 99909 [3]. Hence, the initial increment is 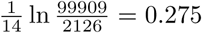. Given the known probability of mutation *m*=0.39 and a value of the parameter for the rate of exit from the disease *γ* = 0.05, this, in accordance with the relation *α* = (1 – *m*) *β – γ*, gives an estimate of the initial virus spread rate at the start of the epidemic: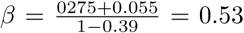. It is this value of that leads to the model graph in the Fig.8.

**FIG. 8.**
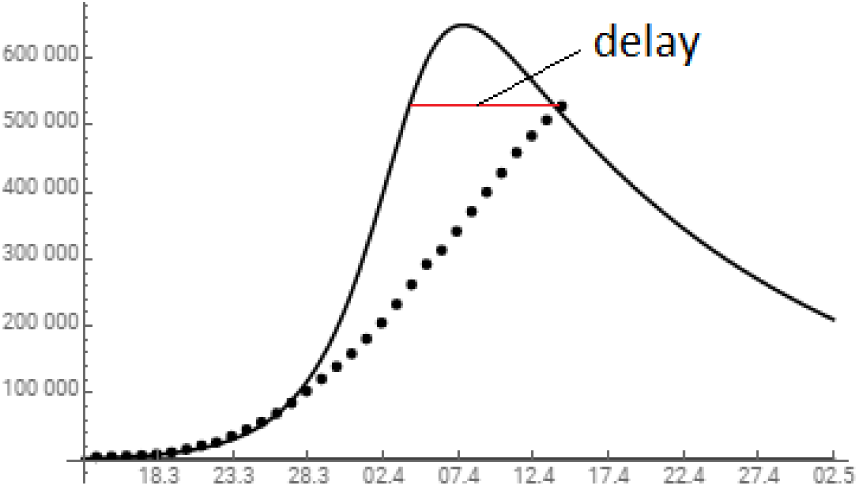
Comparison of the dynamics of the number of cases during the COVID-19 epidemic in USA according to the SIMR model (solid line) with statistical data [3] (dots). The SIMR parameters are taken as *m*=0.39, *β*=0.53, *γ*=0.05.

The dimensionless epidemic factor from here is *r* = (1 – *m*) *β/γ* = 6.5. Thus, the epidemic condition *r >* 1 is fulfilled, as it should be.

The lower limit of hidden immunization in USA in accordance with Eq.(10) is currently about 9%.

It is clearly seen that after the first week of the epidemic, the growth in the number of sick people slowed noticeably compared to the SIMR model with a constant value of *β*. It is natural to consider this as a consequence of the restrictive measures taken, leading to the decrease of *β*.

Comparison of the theoretical and observed dependencies *I*(*t*) shows a lag of the latter by about 8 days. This indicates a later onset of the peak compared with the case of constant *β*, and the delay scale has already been determined, although it may slightly increase. Further, it is natural to assume that restrictive measures can only reduce the height of the maximum *I*(*t*), which for the theoretical curve is at the level of 600 thousand.

### F. South Korea

In South Korea, as in other countries of Southeast Asia, the epidemic developed much more slowly than in the West. The data of South Korea, presented in Fig. 9, are very different from the data of the South of Europe and the USA. At the maximum of the epidemic, the number of infected people did not reach here even 8 thousand. To understand the reason for this difference, two facts must be taken into account: 1) the geographical proximity of South Korea and Wuhan, and 2) the epidemic in South Korea began in mid-February, when it was at its peak in Wuhan. As shown earlier in section II, the red line in Fig.3, the hidden immunization in Wuhan at this point was 90%. This means that 90% of virus came in South Korea from Wuhan already mutated. This was exactly the composition of the virus population got in South Korea from Wuhan.

Accordingly, it is necessary to introduce this fact into the initial conditions of infection in South Korea: instead of the initial condition *M* (0) = 0 common to all previous cases, here it is necessary to take *M* (0) = 9*I*_0_. This means that the hidden immunization with mutated virus from Wuhan has been taking place in South Korea since the very start of the epidemic.

The initial value of the increment is obtained from the number of sick people on 22.02.2020, *I*(0) = 416 and after 10 days - on 03.03.2020, *I*(10) = 5120 [3]. Hence, the initial increment is 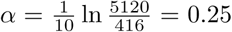. Given the known probability of mutation *m*=0.39 and a value of the parameter for the rate of exit from the disease *γ* = 0.03, this, in accordance with the relation *α* = (1 – *m*) *β – γ*, gives an estimate of the initial virus spread rate at the start of the epidemic: 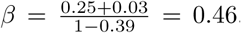. It is this value of *β* that leads to the model graph in the Fig.9.

**FIG. 9.**
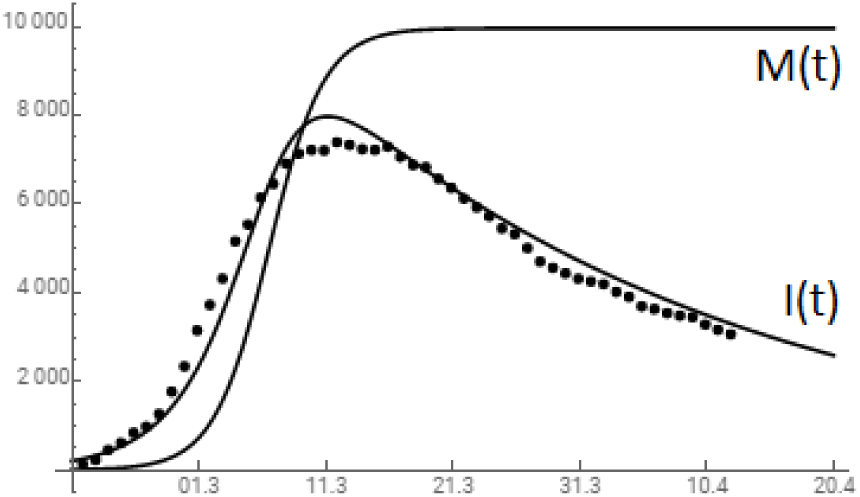
Comparison of the dynamics of the number of cases during the COVID-19 epidemic in South Korea according to the SIMR model (solid line) with statistical data [3] (dots). The SIMR parameters are taken as *m*=0.39, *β*=0.46, *γ*=0.03.Saturation of *M* (*t*) corresponds to 100% hidden immunization. Fraction of the mutated virus in the total initial infection is taken to be *f* =0.9.

It is important to understand that South Korea did not go through one, but two epidemics at once - the initial aggressive and the new mutated virus. The same SIMR model shows that if South Korea entered the epidemic along with Wuhan and got only the original strain of the virus, the number of cases at the peak of the epidemic would not be 8, but 30 thousand. It is the entry of the mutated virus into the epidemic that explains the SIMR model of a much milder course of the epidemic in the entire region of Southeast Asia.

### G. World

At last, we consider the summary data starting on March 10, which we will conditionally consider as the beginning of the post-Wuhan pandemic. Let’s combine them with the result of the SIMR model, as shown in Fig. 10.

**FIG. 10.**
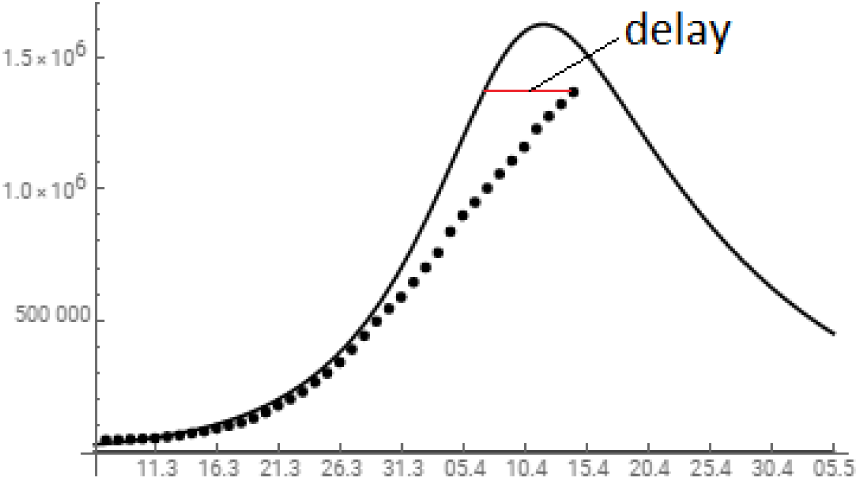
Comparison of the dynamics of the number of cases during the COVID-19 epidemic in the world according to the SIMR model (solid line) with statistical data [3] (dots). The SIMR parameters are taken as *m*=0.39, *β*=0.32, *γ*=0.065.

The initial value of the increment is obtained from the number of sick people on 10.03.2020, *I*(0) = 48031 and after 10 days - on 20.03.2020, *I*(10) = 172591 [3]. Hence, the initial increment is 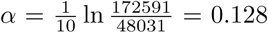. Given the known probability of mutation *m*=0.39 and a value of the parameter for the rate of exit from the disease *γ* = 0.03, this, in accordance with the relation *α* = (1 – *m*) *β – γ*, gives an estimate of the initial virus spread rate at the start of the epidemic: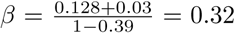. It is this value of *β* that leads to the model graph in the Fig.10.

The dimensionless epidemic factor from here is *r* = (1 – *m*) *β/γ* = 3.0. Thus, the epidemic condition *r >* 1 is fulfilled, as it should be.

It is clearly seen that after the first week of the epidemic, the growth in the number of sick people slowed noticeably compared to the SIMR model with a constant value of *β*. It is natural to consider this as a consequence of the restrictive measures taken, leading to the decrease of *β*.

A comparison of the theoretical and observed dependencies *I*(*t*) shows a noticeable lag of the latter by about 3 days. This indicates a later onset of the peak compared with the case of constant *β*, and the delay scale has already been determined, although it may slightly increase. Further, it is natural to assume that restrictive measures can only reduce the height of the maximum *I*(*t*), which for the theoretical curve is at the level of 1,500 thousand.

## V. CONCLUSIONS

The SIMR model constructed in our work is a direct generalization of the basic SIR model that takes into account the accelerated evolution of the virus during an epidemic within the framework of the simplest approximation of two strains. However, it has the same limitations as the base model. It does not take into account two circumstances: 1) heterogeneity of parameters even within one country, and 2) the age structure in the population.

The first of them should lead to territorial heterogeneity of the rate of spread of the virus, *β*, which, apparently, increases with increasing population density. We smooth out this factor by considering the reference regions with the highest concentration of the population.

The second is related to the age-dependent level of immunity, which determines the rate of at which the body emerges from the disease. This requires a separate consideration, going beyond the framework of the constructed simple SIMR model. At the same time, even it allows one to qualitatively assess the effect of the average age of the population on the course of the epidemic. In countries with a predominantly elderly population (Italy), there is a longer course of the epidemic with a higher gentle maximum and a longer exit from it. This is due precisely to the low value of the average immunity parameter *γ* in elderly patients.

We emphasize once again that the main property of the solutions of the SIMR model is the avalanche-like spread of the mutated strain, which is faster than the epidemic itself. The hidden immunization of the population resulting from this spread stops the epidemic.

Now we are in a position to evaluate the effect of all three model parameters on the incidence curve *I*(*t*) in an epidemic. It always begins with an ascent, then turning to a maximum and a descent.

The steepness of the rise is determined by the spread rate, *β*. This is exactly the parameter that is influenced by quarantine measures.

The height of the maximum in the SIMR model is determined by the virus mutation factor, m. It is impossible to influence this factor, but knowing that it is possible to forecast the course of the epidemic, which allows us to plan and allocate resources. We consider this parameter as inherent property of the virus, a kind of epidemic index, which determines its extent.

The steepness of the descent depends on the *γ*-parameter that is on the rate of gaining immunity of the sick persons and - to some extent - on the quality of medical care.

The experience of the Wuhan epidemic shows that it is quarantine measures that can have a significant - albeit limited - impact on the height of the maximum epidemic. In this particular case, according to our model, it was reduced by about 1/3, i.e. for 25 thousand infected. Accordingly, with an average mortality rate of 6%, this saved approximately 1,500 lives.

In general, the development of a pandemic has a limited time frame, practically independent of efforts in a particular country. However, the height of the maximum epidemic in each country is determined by the intensity of timely quarantine measures. This reduces the steepness of the rise and allows to go through the epidemic without rising to its top, but to make it in a “tunnel” way. As it was in China and, apparently, in Germany (see Fig.3 and Fig.6).

## Data Availability

used open data only

## ACKNOWLEDGEMENT

We are indebted to Prof. Mykola Iabluchanskiy for valuable remarks and consultations.

## Appendix A

It is convenient to explore the role of mutation factor *m* in the absence of immunity (*γ* = 0, *R* = 0). We choose a unit of time so that *β* = 1. Then the SIMR model turns into an exactly solvable three-component SIM model

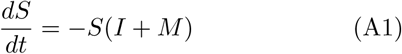

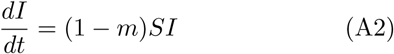

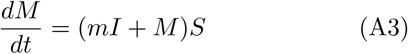

keeping the sum *S* + *I* + *M* = 1. The equation for *S* is separated:

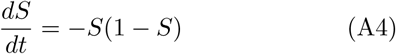

and has a solution

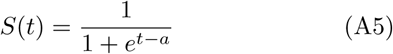

The initial conditions *I*(0) = *I*_0_ and *S*(0) = 1 – *I*_0_ fix the value of parameter *a* by the relation

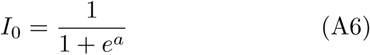

After that, the second equation for *I*(*t*) is integrated:

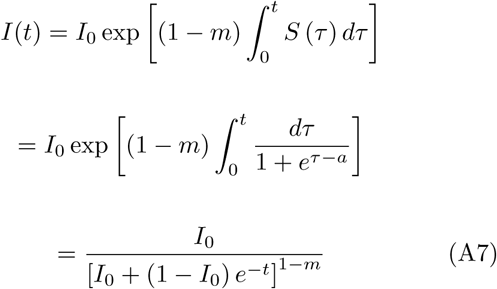

Of interest is the asymptotic behaviour of the fraction of sick people *I*(*t*) at *t* → ∞, that is expressed as

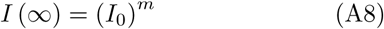

It is Eq.(9) of the main text. For example, for *I*_0_ = 10^*-*6^ we have for the asymptotic of the sick people fraction *I* (∞)

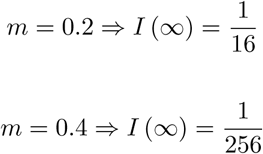

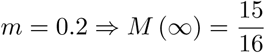

Accordingly, the asymptotic of the hidden immunization *M* (∞) = 1 *-I* (∞) in the same cases are

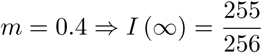

These examples show how the epidemic outcome changes with increasing mutation parameter *m*.

Note that the found relation (A8) holds for any monotonic dependence *β*(*t*) corresponding to a change in the transmission rate *β* during the course of the epidemic due to quarantine restrictions. The only condition is the fulfilment of the relation *r* = (1 − *m*) *β/γ ≫* 1, which allows neglecting the immunity factor *γ*, as done in equations (A1-A3).

This allows to express the hidden immunization *M* through the fraction of sick people*I*independently on the concrete function*β*(*t*). Dividing Eq.(A3) through Eq.(A2) results in following uniform differential equation

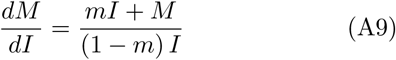

This equation for unknown function *M* (*I*) has simple solution

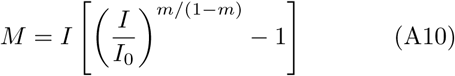

which corresponds to initial condition *M* (*I*_0_) = 0, that is at *t* = 0 we have initial fraction of sick people *I*(0) = *I*_0_ and no hidden immunization *M* (0) = 0.

As this relation is universal for *r ≫* 1, and numbers of sick people *I*_0_, *I* are always known, it allows to trace the level of the hidden immunization at any time.

## Appendix B

Below we present the code for Mathematika, which solves the system of equations of the SIMR model (5-8) and represents the solution in the form of a graph.

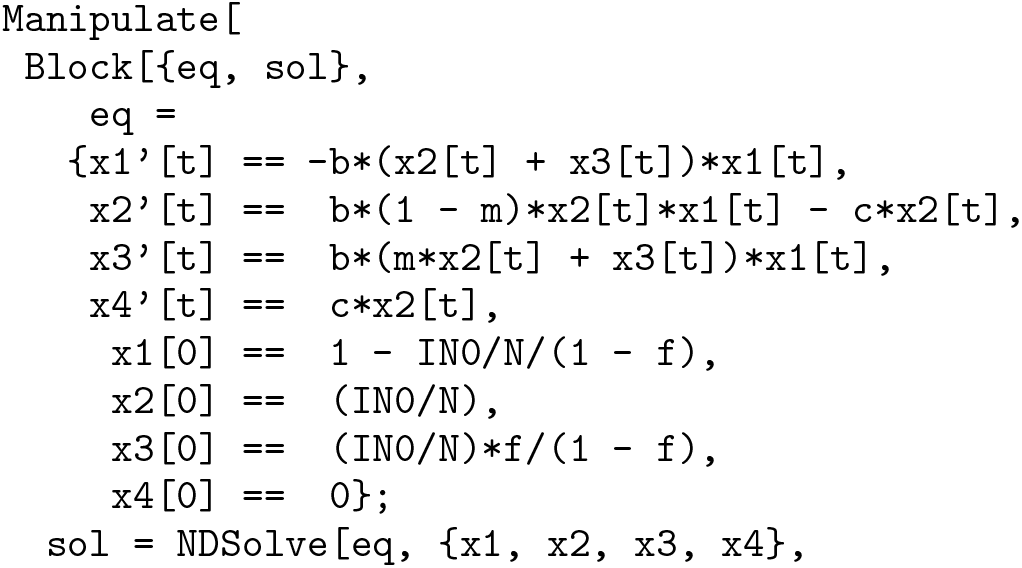
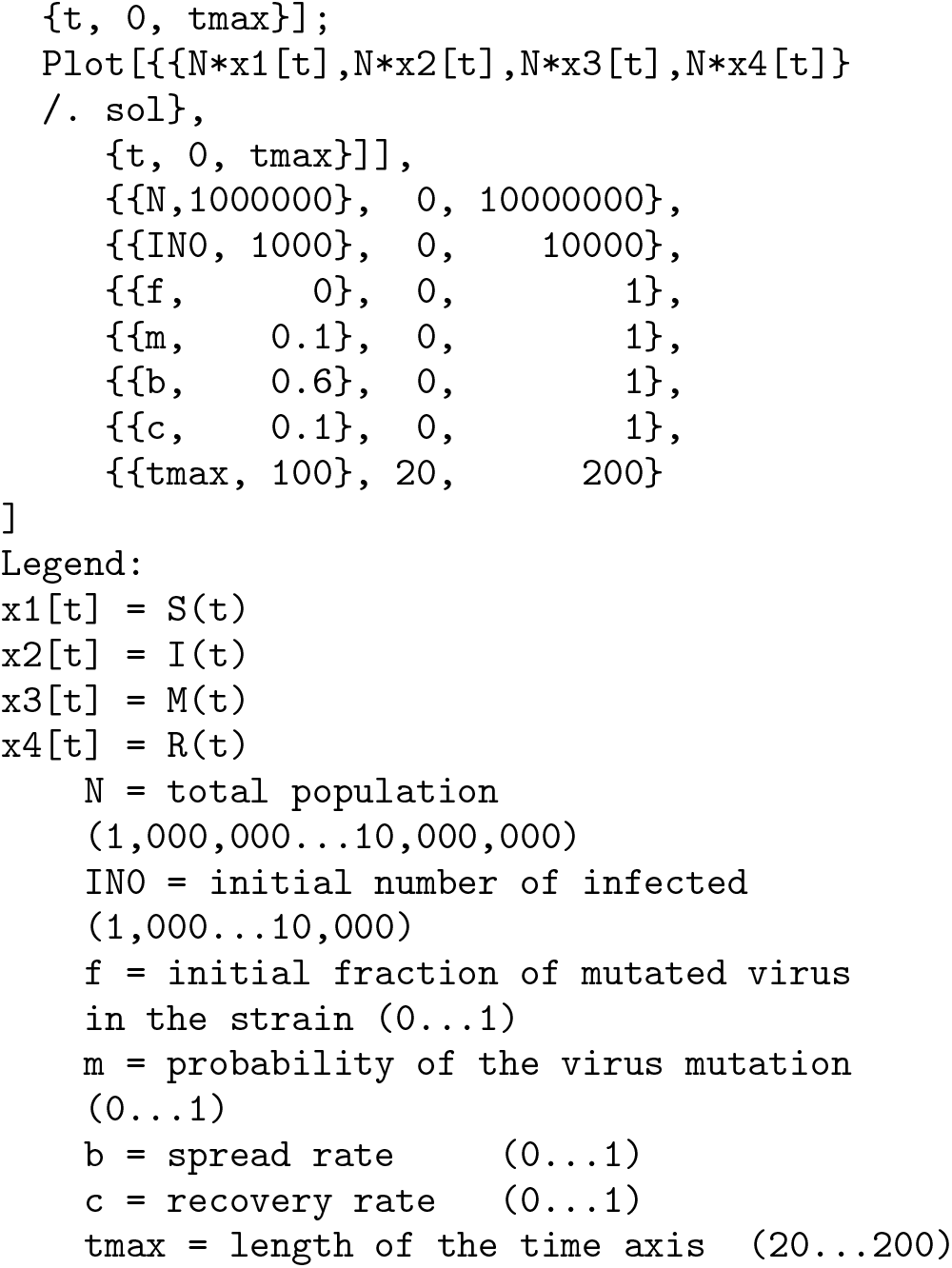

## Notes

### Competing Interest Statement

The authors have declared no competing interest.

### Funding Statement

no funding support

## REFERENCES

[1] Kermack, W. O. and McKendrick, A. G. “A Contribution to the Mathematical Theory of Epidemics.” Proc. Roy. Soc. Lond. A 115, 700–721, 1927.

2. Anderson, R. M. and May, R. M. “Population Biology of Infectious Diseases: Part I.” Nature 280, 361–367, 1979.

3. Harko, Tiberiu; Lobo, Francisco S. N.; Mak, M. K. “Exact analytical solutions of the Susceptible-Infected-Recovered (SIR) epidemic model and of the SIR model with equal death and birth rates”. Applied Mathematics and Computation. 236, 184194, 2014.

4. https://mathworld.wolfram.com/Kermack-McKendrickModel.html

[2] https://www.gisaid.org/epiflu-applications/next-hcov-19-app/

[3] https://www.worldometers.info/coronavirus/

[4] https://www.dropbox.com/s/oxmu2rwsnhi9j9c/Draft-COVID-19-Model%20%2813%29.pdf?dl=0

